# Cytoplasmic Expression of LMP1 and its Nuclear Translocation in Nasopharyngeal Carcinoma Correlates with Epithelial-mesenchymal Transition

**DOI:** 10.1101/2022.02.13.22270759

**Authors:** Weiren Luo

## Abstract

Up to now, the prognostic significance of latent membrane protein 1 (LMP1) in nasopharyngeal carcinoma (NPC) tissues still remains controversial. This study aims to investigate aberrant localization of LMP1 and its relationship with epithelial-mesenchymal transition (EMT) in NPC samples by immunohistochemistry and immunofluorescence. Both cytoplasmic and nuclear LMP1 expressions were observed in NPC tissues. In some tissues, nuclear LMP1 was frequently observed at tumor invasive front and tumor buddings. Nuclear LMP1 expression was significantly associated with lymph node metastasis (*P*=0.031), local recurrence (*P*=0.002), lymphatic invasion (*P*= 0.004) and tumor budding (*P*=0.001). Furthermore, nuclear LMP1 showed significant correlations with EMT markers including E-cadherin (*P*=0.037), Vimentin (*P* < 0.001), N-cadherin (*P*=0.003), Snail (*P*=0.003) and Twist (*P*=0.002), but not significantly linked with Fibronectin (*P*=0.103) and Slug (*P*=0.503). According to cytoplasmic LMP1, it correlated strongly with lymphatic invasion (*P*=0.044), vascular invasion (*P*=0.003) and EMT proteins including E-cadherin (*P*=0.014), Vimentin (*P=*0.006), N-cadherin (*P*=0.003), Snail (*P*=0.008) and Slug (*P*=0.007), whereas not significantly associated with Fibronectin (*P*=0.221) and Twist (*P*=0.106). However, multivariate analysis showed that nuclear LMP1 (*P*=0.844) and cytoplasmic LMP1 (*P*=0.291) were not independent predictors for NPC. In conclusion, we demonstrate firstly that abnormal localization of LMP1 correlates with EMT properties and aggressiveness in NPC, respectively.

## Introduction

Nasopharyngeal carcinoma (NPC) is the most common type of the head and neck cancers in southern China and is strongly associated with Epstein-Barr virus (EBV) infection (Wong KCW *et al*. 2021; Luo *et al*. 2013). Latent membrane protein 1 (LMP1), is the major EBV-encoded protein essential for EBV-induced B cell transformation and immune surveillance (Yoshizaki *et al*. 2013). *In vitro*, LMP1 could promote cell invasion, and metastasis of NPC, indicating that interfering LMP1 pathways could be a promising anti-therapy strategy (Dawson *et al*. 2012). However, the prognostic significance of LMP1 in NPC tissues remains controversial. Several reports have showed that LMP-1 expression contributed to poor prognosis of patients (Hariwiyanto *et al*. 2010; Sarac *et al*. 2001; Taheri-Kadkhoda *et al*. 2009). In contrast, other studies showed the opposite conclusions (Plaza *et al*. 2003; Li *et al*. 2009; Chen *et al*. 2006). Generally, LMP1 protein is found in the cytoplasm (or membrane) of cancer cells. On the other side, several observations have also revealed that LMP1 was localized in the nucleus of human cells. For example, Xu J et al reported that the expression of LMP-1 protein was preferentially observed in the nucleus of human T cells and induced the malignant transformation of EBV genome-positive T-Cell (Xu *et al*. 2002). However, the localization of nuclear LMP1 and its prognostic significance in NPC remain largely unknown.

Epithelial-mesenchymal transition (EMT) is the key process for cancer invasion, metastasis in various types of human cancers (Brabletz *et al*. 2018; Zidar *et al*. 2018). It has also been reported that EMT contributed to tumor metastasis in NPC and was induced by ectopic expression of LMP1 *in vitro* (Horikawa *et al*. 2007; Horikawa *et al*. 2011; Shair *et al*. 2009). Ten years ago, we demonstrated that neoplastic spindle cells possessing features of EMT should be considered as the more aggressive subtype in NPC, and EBER and LMP1 were highly expressed in these spindle cells (Luo *et al*. 2012a; Luo *et al*. 2013). However, as far as we know, the association between different localization of LMP1 protein and EMT in cancer tissues has not been fully described.

Therefore, the aim of this study was to clarify whether the different localization of LMP1 has clinicopathological impacts on NPC progression. In particular, the relationship between aberrant expression of LMP1 protein and EMT-related biomarkers such as E-cadherin, Vimentin, N-cadherin and Snail in NPC was also examined.

## Material and methods

### Patients and samples

In total, 136 formalin-fixed paraffin-embedded tissues diagnosed with NPC during the period of 2003 to 2007 were obtained from the Department of Pathology, Guangdong Medical College, China. None of the patients had received therapy before. This protocol was approved by the Ethics Committee of Guangdong Medical College. Histopathologic classification of primary NPC samples was done according to “the Pathology and genetics of head and neck tumours” by the World Health Organization (WHO). All of the tumors were classified as non-keratinizing NPC. Among them, 26 samples were differentiated non-keratinizing carcinomas (DNKC) and 110 samples were undifferentiatied carcinomas (UDC). Clinical stage of these patients was classified based on the Tumor-Node-Metastasis (pTNM) system (AJCC/UICC). The clinicopathological characteristics of NPC patients were summarized in **Supplemental Table 1**. In the present study, the end date of the follow-up study was August 2012. The patients were followed-up from 8 to 106 months, with a mean period of 65.0 months. In addition, 45 non-tumoral pharynx tissues were obtained as normal controls.

### Tissue microarray construction

After the cancerous and non-neoplastic pharynx epithelium areas of H&E-stained arrays were carefully reviewed and determined, tissue microarray (TMA) was built as described previously (Luo *et al*. 2013). In brief, representative areas of the paraffin ‘donor’ blocks were marked on the slide. Two tissue cylinders with a diameter of 1.5 mm were taken from 136 NPC tissues and 45 non-cancerous nasopharyngeal tissues and precisely arrayed into new paraffin blocks using a TMA workstation (Beecher Instruments, Silver Spring, MD, USA). All patients were represented by two cancer cores.

### Immunohistochemistry and immunofluorescence

The sections were put into in citrate buffer (PH 6.0) for high-pressure antigenic retrieval and boiled for 2 minutes. The endogenous peroxidase activity and non-specific binding were eliminated by 3% hydrogen peroxide (H2O2) and 1% bovine serum albumin (BSA) at room temperature, respectively. The sections were incubated overnight at 4°C with the primary antibodies. The primary antibodies were listed as follows: LMP1 (Santa Cruz, clone CS1/2/3/4, dilution 1:25); E-cadherin (BD, clone 36/E, dilution 1:300); N-cadherin (Abcam, clone 5D5, dilution 1:300); Vimentin (BD, clone RV202, dilution 1:200); Fibronectin (Abcam, clone IST-9, dilution 1:300); Snail (Cell Signaling, clone C15D3, dilution 1:100); Snail (Cell Signaling, clone C15D3, dilution 1:100); Slug (Cell Signaling, clone C19G7, dilution 1:50); Twist (Abcam, clone 3E10, dilution 1:400); D2-40 (Zymed, clone D2-40, dilution 1:200); CD31 (Zymed, clone JC70A, dilution 1:200); Pan-cytokeratin (Zymed, clone AE1/AE3, dilution 1:250). The slides were then washed with PBS for 2 × 5 min and incubated with the biotinylated secondary antibody (Zymed, San Francisco, CA) and streptavidin horseradish peroxidase complex for 15 min at room temperature, respectively. Subsequently, sections were reacted with 3,3′-diaminobenzidine (DAB) for 2 min for visualization, and counterstained with hematoxylin. PBS was used instead of the primary antibodies as negative controls.

The protocol of immunofluorescence labelling of formalin-fixed (FFPE) was equal to IHC procedure before sections of cancer tissues were incubated with primary antibodies over night at 4°C. After washing two times in PBS for 10 min, these sections were stained with Dylight594-conjugated goat anti-mouse antibody (Jackson, dilution 1:600) for 2 h at room temperature. At last, the slides were reacted with DAPI (Sigma) for 10 min and mounted in Antifade Medium (P0126, Beyotime).

### Evaluation of immunostaining

Evaluation of the immunostaining expression was examined separately by two pathologists who were unaware of the clinical data. Four views were examined per case, and at least 400 cancer cells were counted per view at ×400 magnification. As described previously, the immunohistochemical staining was graded on the basis of the intensity and percentage of positive cancer cells (Luo *et al*. 2013; Li *et al*. 2008). With regard to the scoring criteria for intensity, it was recorded on a scale of 0 to 4 as follows: 0 (absence of reactivity), 1 (weak reactivity), 2 (moderate reactivity), and 3 (strong reactivity). Relative to the percentage of positive tumor cells, it was reviewed as follows: 0 (no positive cells), 1 (<10% positive cells), 2 (10-50% positive cells), and 3 (>50% positive cells).

The expression patterns of proteins were calculated by multiplying the scores of staining intensity and the proportion of positive tumor cells. Taken together, a score of ≤ 4 was regarded as patients with low expression and ≥6 as those with high expression in tissue sections. In addition, D2-40 and CD31was used to confirmed tumor cells with lymphatic invasion or vascular invasion, respectively. The numbers of tumor budding cells was stained and mounted by Pan-cytokeratin (budding <5 and ≥ 5 was recognized as low and high grade, respectively) (Luo *et al*. 2012; Brown *et al*. 2010).

### Statistical analysis

All statistical analyses were performed using the SPSS13.0 (SPSS Inc., Chicago, IL) statistical software package. Comparisons between LMP1/EMT proteins and clinicopathologic parameters were done with the χ2 test. Spearman correlation coefficient was used to evaluate the relationships between variables. Survival curves were plotted by the Kaplan-Meier method and the log-rank test. A Cox regression (Proportional hazard model) was used for univariate and multivariate analysis. The “*” sign denotes *P* < 0.05 compared with control and the “**” sign denotes P < 0.01.

## Results

### Cytoplasmic and nuclear LMP1 is overexpressed in NPC samples

Immunohistochemical results were summarized in **Supplemental Table 1**. LMP1 protein was weakly expressed in the non-cancerous nasopharyngeal epithelium (Fig. 1A, 1B), only 2 of 45 cases (4.4%) showed high expression. On the contrary, high expression of LMP1 was found in the cytoplasm and nucleus of tumor cells. Among 136 NPC biopsies, cytoplasmic and nuclear of LMP1 overexpression was detected in 30 cases (22.1%, Fig. 1D, 1E) and 51 cases (37.5%, Fig.1G, 1H), respectively. Furthermore, cytoplasmic (Fig. 1F) and nuclear localization of LMP1 (Fig. 1I) was confirmed by immunofluorescence labeling in NPC tissues.

**FIG. 1.**
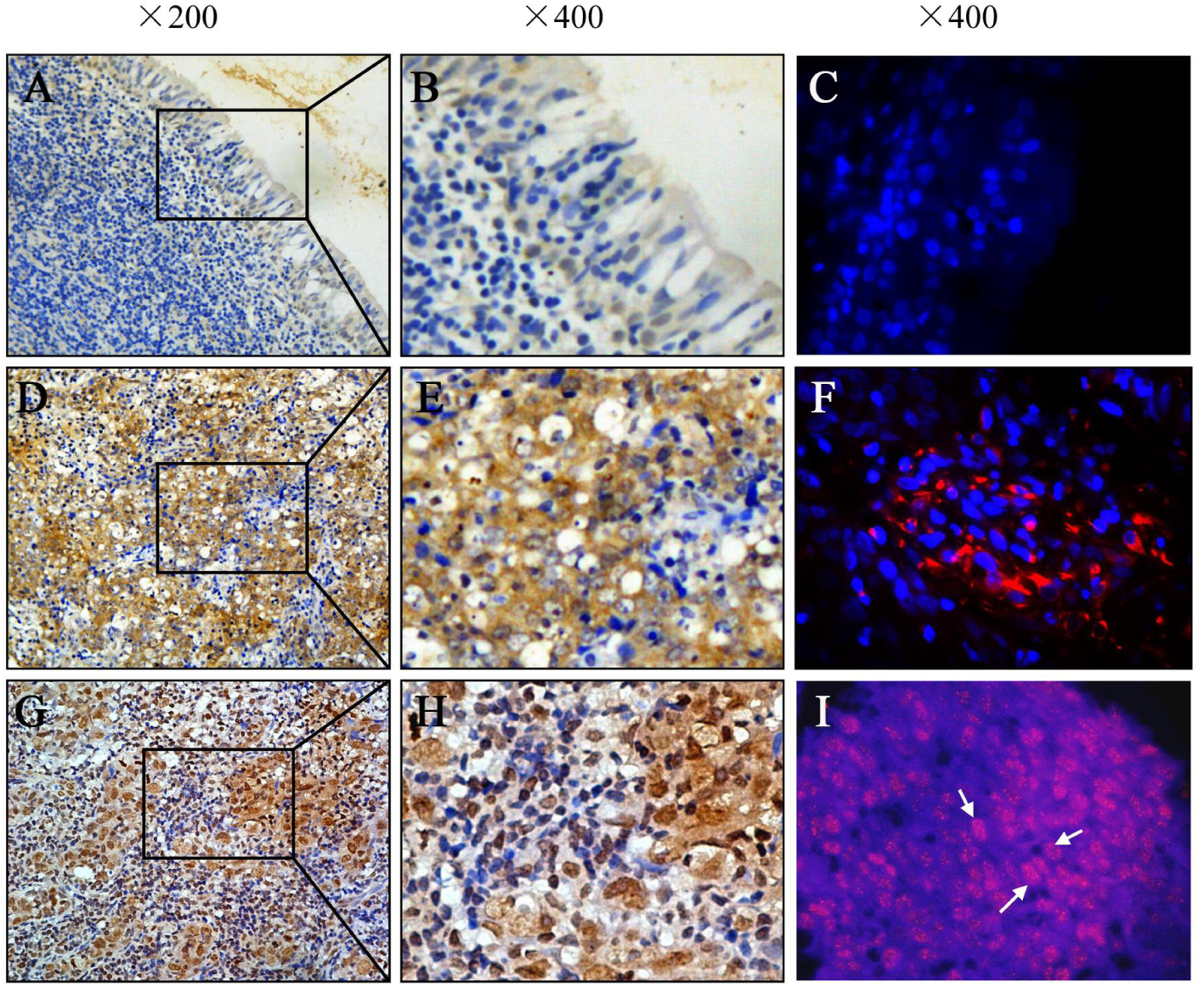
The expression and localization of LMP1 in paraffin-embedded nasopharyngeal carcinoma (NPC) tissues. LMP1 was weakly expressed in non-tumoral epithelium (**A, B**). Immunofluorescence showed that LMP1 was negative in normal epithelium (**C**). High expression of cytoplasmic LMP1 (**D, E**) were observed in NPC. Immunofluorescent labeling of LMP1 showed cytoplasmic localization (red) of tumor cells (**F**). DAPI (blue). High nuclear expression of LMP1 was detected in NPC (**G, H**). Immunofluorescent labeling of nuclear localization of LMP1 (red) in tumor cells (**I**).

### Association of abnormal LMP1 with clinicopathological variables in NPC

The relationship of different localization of LMP1 expression and clinicopathological parameters was depicted in **Table 1** and Fig. 2. High expression of cytoplasmic LMP1 was positively associated with lymphatic invasion (*P* = 0.044) and vascular invasion (*P* = 0.003). However, no significant association was identified between cytoplasmic LMP1 and other variables including gender (*P* =0.271), age (*P* = 0.215), histologic type (*P*=0.699), T classification (*P*=0.486), lymph node metastasis (*P*=0.266), distant metastasis (*P*= 0.434), clinical stage (*P* = 0.272), local recurrence (*P* = 0.537) and tumor budding (*P*=0.489). As shown in Fig. 2G, the expression of nuclear LMP1 was significantly associated with lymph node metastasis (*P*= 0.031), local recurrence (*P*= 0.002), lymphatic invasion (*P*= 0.004) and tumor budding (*P*= 0.001). High expression of total LMP1 correlated significantly with several clinicopathological variables including T classification (*P*= 0.037), lymphatic invasion (*P* = 0.001), lymph node metastasis (*P*= 0.039) and tumor budding (*P* = 0.001) in patients.

**Table 1.** Relationship between aberrant expression levels of LMP1 protein and clinicopathological variables in 136 NPC patients

| *Variables* | *Cases* | *Cytoplasmic LMP1 (n, %)* |  |  | *Nuclear LMP1 (n, %)* |  |  | *Invasive front nuclear LMP1 (n, %)* |  |  |
| --- | --- | --- | --- | --- | --- | --- | --- | --- | --- | --- |
|  |  | *Low* | *High* | *P\** | *Low* | *High* | *P\** | *Low* | *High* | *P\** |
| Gender |  |  |  |  |  |  |  |  |  |  |
| Women | 33 | 28 (84.8) | 5 (15.2) | 0.271 | 22 (66.7) | 11 (33.3) | 0.570 | 29 (82.5) | 4 (17.5) | 0.467 |
| Men | 103 | 78 (75.7) | 25 (24.3) |  | 63 (61.2) | 40 (38.3) |  | 85 (87.9) | 18 (12.1) |  |
| Age (y) |  |  |  |  |  |  |  |  |  |  |
| <48 | 68 | 56 (82.4) | 12 (17.6) | 0.215 | 47 (69.1) | 21 (30.9) | 0.111 | 59 (86.8) | 9 (13.2) | 0.352 |
| ≥48 | 68 | 50 (73.5) | 18 (26.5) |  | 38 (55.9) | 30 (44.1) |  | 55 (80.9) | 13 (19.1) |  |
| Histologic type |  |  |  |  |  |  |  |  |  |  |
| DNKC | 26 | 21 (80.8) | 5 (19.2) | 0.699 | 17 (65.4) | 9 (34.6) | 0.735 | 19 (73.1) | 7 (26.9) | 0.098 |
| UDC | 110 | 85 (77.3) | 25 (22.7) |  | 68 (61.8) | 42 (38.2) |  | 95 (86.4) | 15 (13.6) |  |
| T classification |  |  |  |  |  |  |  |  |  |  |
| T1-T2 | 62 | 50 (80.6) | 12 (19.4) | 0.486 | 44 (71.0) | 18 (29.0) | 0.062 | 57 (91.9) | 5 (8.1) | 0.019 |
| T3-T4 | 74 | 56 (75.7) | 18 (24.3) |  | 41 (55.4) | 33 (44.6) |  | 57 (77.0) | 17 (23.0) |  |
| Lymph node metastasis |  |  |  |  |  |  |  |  |  |  |
| N0-N1 | 80 | 65 (81.3) | 15 (18.7) | 0.266 | 56 (70.0) | 24 (30.0) | 0.031 | 74 (92.5) | 6 (7.5) | 0.001 |
| N2-N3 | 56 | 41 (73.2) | 15 (26.8) |  | 29 (51.8) | 27 (48.2) |  | 40(71.4) | 16 (28.6) |  |
| Distant metastasis |  |  |  |  |  |  |  |  |  |  |
| No | 115 | 91 (79.1) | 24 (20.9) | 0.434 | 75 (65.2) | 40 (34.8) | 0.126 | 98 (85.2) | 17 (14.8) | 0.302 |
| Yes | 21 | 15 (71.4) | 6 (28.6) |  | 10 (47.6) | 11 (52.4) |  | 16 (76.2) | 5 (23.8) |  |
| Clinical stage |  |  |  |  |  |  |  |  |  |  |
| I - II | 38 | 32 (84.2) | 6 (15.8) | 0.272 | 28 (73.7) | 10 (26.3) | 0.093 | 35 (92.1) | 3 (7.9) | 0.102 |
| III-IV | 98 | 74 (75.5) | 24 (24.5) |  | 57 (58.2) | 41 (41.8) |  | 79 (80.6) | 19 (19.4) |  |
| Local recurrence |  |  |  |  |  |  |  |  |  |  |
| No | 103 | 79 (76.7) | 24 (23.3) | 0.537 | 72 (69.9) | 31 (30.1) | 0.002 | 91 (88.3) | 12 (11.7) | 0.011 |
| Yes | 33 | 27 (81.8) | 6 (18.2) |  | 13 (39.4) | 20 (60.6) |  | 23 (69.7) | 10 (30.3) |  |
| Lymphatic invasion |  |  |  |  |  |  |  |  |  |  |
| Negative | 97 | 80 (82.5) | 17 (17.5) | 0.044 | 68 (70.1) | 29 (29.9) | 0.004 | 86 (88.7) | 11 (11.3) | 0.016 |
| Positive | 39 | 26 (66.7) | 13 (33.3) |  | 17 (43.6) | 22 (56.4) |  | 28 (71.8) | 11 (28.2) |  |
| Vascular invasion |  |  |  |  |  |  |  |  |  |  |
| Negative | 108 | 90 (83.3) | 18 (16.7) | 0.003 | 65 (60.2) | 43 (39.8) | 0.273 | 90 (83.3) | 18 (16.7) | 0.760 |
| Positive | 28 | 16 (57.1) | 12 (42.9) |  | 20 (71.4) | 8 (28.6) |  | 24 (85.7) | 4 (14.3) |  |
| Tumor budding |  |  |  |  |  |  |  |  |  |  |
| <5 | 80 | 64 (80.0) | 16 (20.0) | 0.489 | 59 (73.8) | 21 (26.3) | 0.001 | 72 (90.0) | 8 (10.0) | 0.019 |
| ≥ 5 | 56 | 42 (75.0) | 14 (25.0) |  | 26 (46.4) | 30 (53.6) |  | 42 (75.0) | 14 (25.0) |  |

**FIG. 2.**
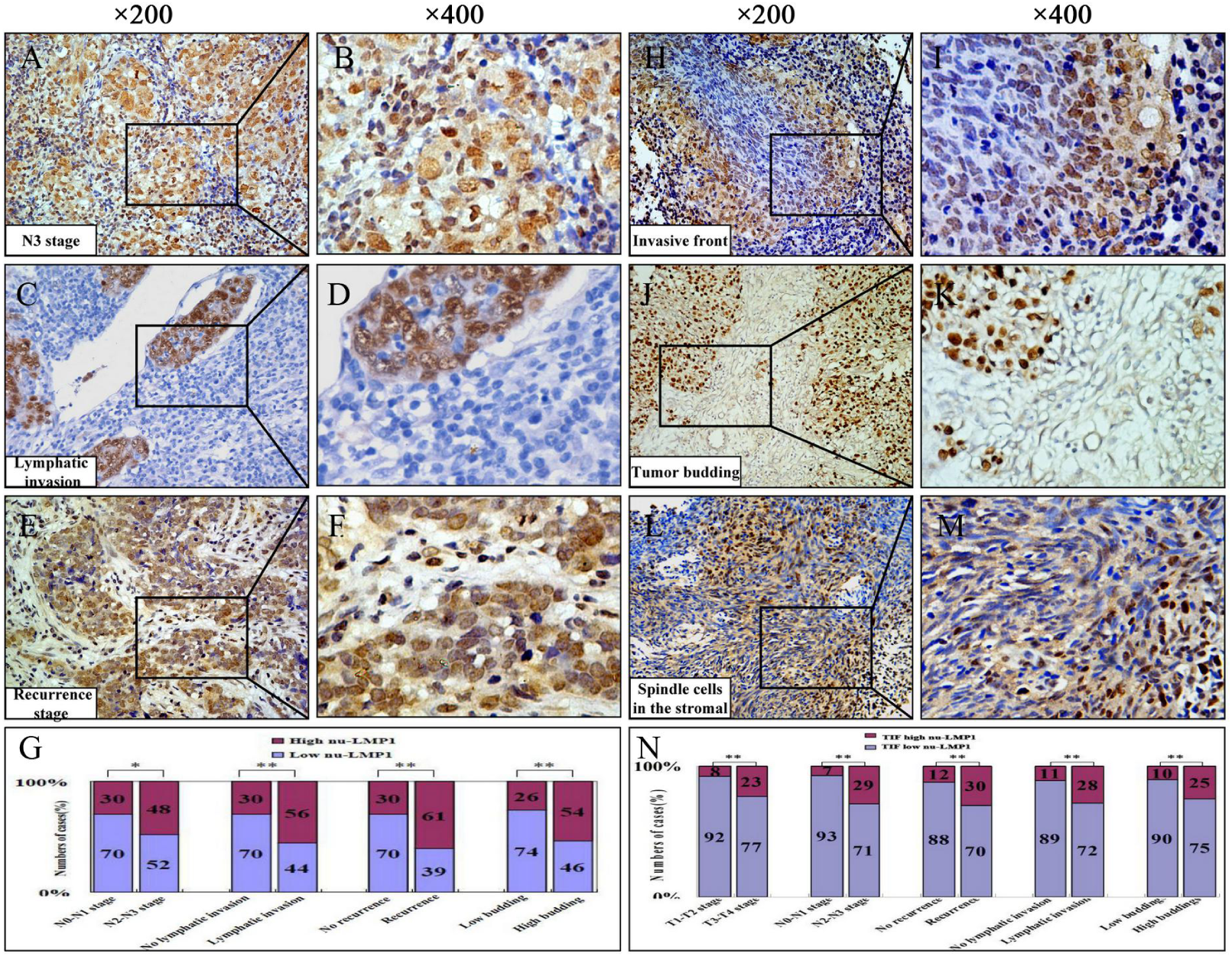
Relationship between nuclear LMP1 expression and clincalpathological features in NPC biopsies. Representative images of high expression of nuclear LMP1 was observed in the N3 (**A, B**), lymphatic invasion (**C**, **D**), recurrence (**E, F**) stages of NPC biopsies. The percentages of high and low expression of nuclear LMP1 according to different clinicopathological features (**G**). High expression of nuclear LMP1 was prominently found at tumor invasive front (**H, I**), budding cells (**J, K**) and cancer spindle cells migrating into the surroundings (**L, M**). High expression of nuclear LMP1 significatly correlated with different clinicopathological features (**N**).

In some cases, high expression of nuclear LMP1 was more frequently observed at the invasive front of tumors (Fig. 2H, I), budding cells (Fig. 2J, K) disseminating from tumors, and cancer cells migrating into the stroma with spindle-shaped phenotype (Fig. 2L, M). Interestingly, a significant association was found between nuclear LMP1 expression in the invasive front with T classification (*P*= 0.019), lymph node metastasis (*P*= 0.001), local recurrence (*P*= 0.011), lymphatic invasion (*P* = 0.016) and tumor budding (*P* = 0.019) (Fig. 2N).

Spearman correlation analysis was further performed to confirm a significantly positive correlation between cytoplasmic LMP1 expression and lymphatic invasion (r = 0.172; *P* = 0.045) and vascular invasion (r = 0.255; *P* = 0.003). On other hand, there was a statistically significant correlation between nuclear expression of LMP1 and lymph node metastasis (r = 0.185; *P* = 0.031), local recurrence (r = 0.270; *P* = 0.001), lymphatic invasion (r = 0.248; *P* = 0.004) and tumor budding (r = 0.278; *P* = 0.001).

### Relations between LMP1 expression and EMT-related markers in NPC

As shown in **Table 2**, cytoplasmic LMP1 expression was significantly associated with E-cadherin (*P* = 0.014), Vimentin (*P* = 0.006), N-cadherin (*P* = 0.003), Snail (*P* = 0.008) and Slug (*P* = 0.007) in NPC samples. On the other hand, in 136 samples with NPC, there was a close linkage between the expression of nuclear LMP1 and E-cadherin (*P* = 0.037), Vimentin (*P* = 0.000), N-cadherin (*P* = 0.003), Snail (*P* = 0.003) and Twist (*P* = 0.002). As shown in Fig. 3, high expression of cytoplasmic and nuclear LMP1 was associated with low expression of E-cadherin, and high expression of N-cadherin, Vimentin and Snail in the same NPC biopsies, respectively. Taken together, we indicate that tumor cells with high cytoplasmic and nuclear LMP1 in NPC might strongly resemble cells that have undergone an EMT.

**Table 2.** Associations between LMP1 expression and EMT-related proteins in 136 patients with NPC

| Variables | Cases | Cytoplasmic LMP1 (n, %) |  |  | Nuclear LMP1 (n, %) |  |  |
| --- | --- | --- | --- | --- | --- | --- | --- |
|  |  | Low | High | P* | Low | High | P* |
| E-cadherin |  |  |  |  |  |  |  |
| Low expression | 104 | 76 (73.1) | 28 (26.9) | 0.014 | 60 (57.7) | 44 (42.3) | 0.037 |
| High expression | 32 | 30 (93.8) | 2 (6.3) |  | 25 (78.1) | 7 (21.9) |  |
| Vimentin |  |  |  |  |  |  |  |
| Low expression | 71 | 62 (87.3) | 9 (12.7) | 0.006 | 56 (78.9) | 15 (21.1) | 0.000 |
| High expression | 65 | 44 (67.7) | 21 (32.3) |  | 29 (44.6) | 36 (55.4) |  |
| N-cadherin |  |  |  |  |  |  |  |
| Low expression | 73 | 64 (87.7) | 9 (12.3) | 0.003 | 54 (74.0) | 19 (26.0) | 0.003 |
| High expression | 63 | 42 (66.7) | 21 (33.3) |  | 31 (49.2) | 32 (50.8) |  |
| Fibronectin |  |  |  |  |  |  |  |
| Low expression | 94 | 76 (80.9) | 18 (19.1) | 0.221 | 63 (67.0) | 31 (33.0) | 0.103 |
| High expression | 42 | 30 (71.4) | 12 (28.6) |  | 22 (52.4) | 20 (47.6) |  |
| Snail |  |  |  |  |  |  |  |
| Low expression | 70 | 61 (87.1) | 9 (12.9) | 0.008 | 52 (74.3) | 18 (25.7) | 0.003 |
| High expression | 66 | 45 (68.2) | 21 (31.8) |  | 33 (50.0) | 33 (50.0) |  |
| Slug |  |  |  |  |  |  |  |
| Low expression | 77 | 66 (85.7) | 11 (14.3) | 0.007 | 50 (64.9) | 27 (35.1) | 0.503 |
| High expression | 59 | 39 (66.1) | 20 (33.9) |  | 35 (59.3) | 24 (40.7) |  |
| Twist |  |  |  |  |  |  |  |
| Low expression | 63 | 53 (84.1) | 10 (15.9) | 0.106 | 48 (76.2) | 15 (23.8) | 0.002 |
| High expression | 73 | 53 (72.6) | 20 (27.4) |  | 37 (50.7) | 36 (49.3) |  |

\* $P < 0.05$ as statistically significant.
EMT, epithelial-mesenchymal transition

**FIG. 3.**
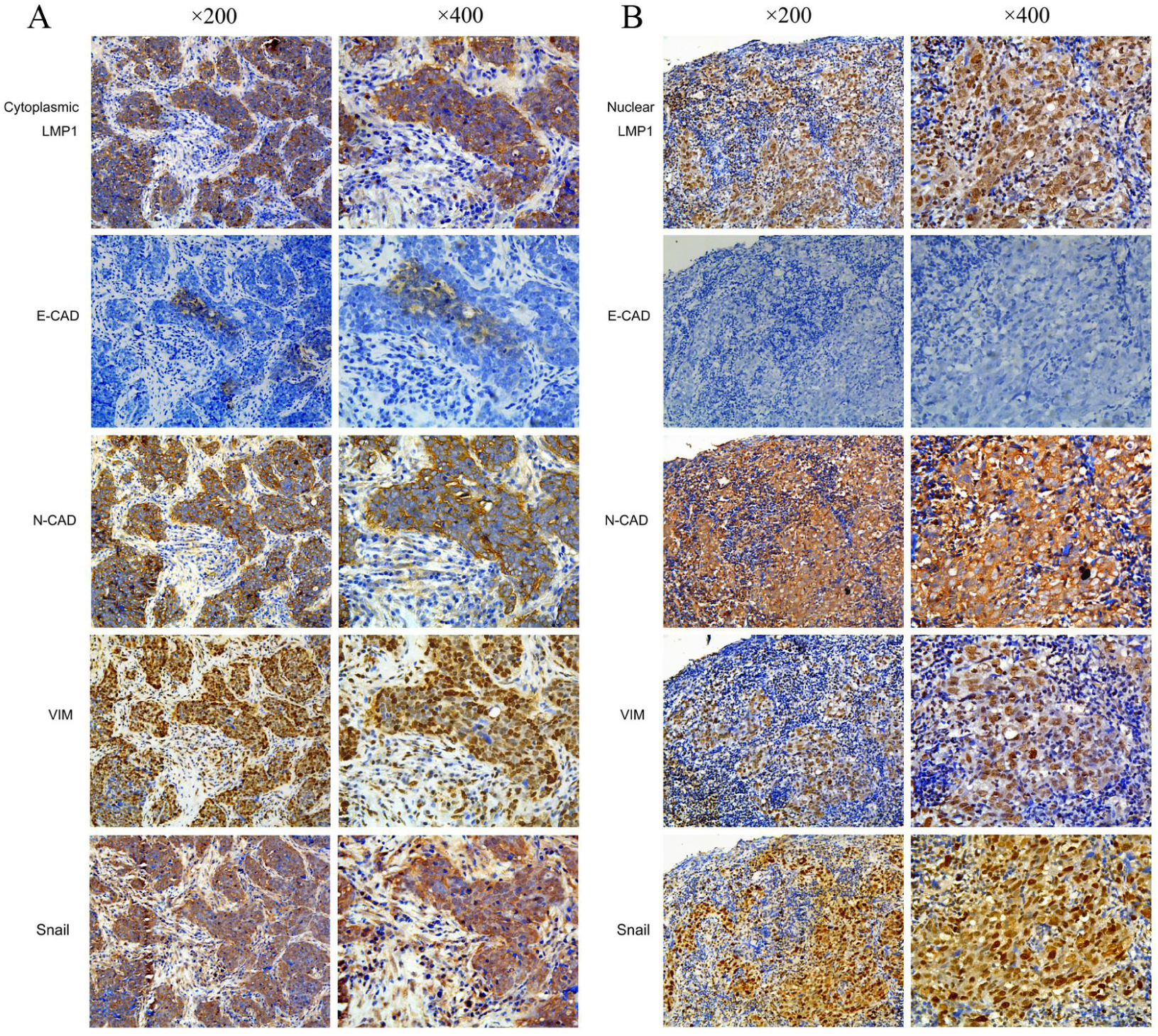
Cytoplasmic and nuclear localization of LMP1 staining correlates strongly with epithelial-mesenchymal transition (EMT) characteristics. Representative images show that high cytoplasmic (**A**) or nuclear expression (**B**) of LMP1 linked negatively with low expression of E-cadherin, whereas positively with high expression of N-cadherin, Vimentin and Snail.

### Association between LMP1 expression and survival

A multivariate Cox regression analysis was performed to determine whether LMP1 patterns have predictive impacts on patients’ clinical outcome (**Table 3**). Distant metastasis (*P*= 0.000), local recurrence (*P*= 0.002) and tumor budding (*P*= 0.003) had independently prognostic values for patient outcome. However, nuclear LMP1 (*P*=0.844), cytoplasmic LMP1 (*P*= 0.291) and other clinicopathologic factors was found not to be independent predictors for NPC.

**Table 3.**
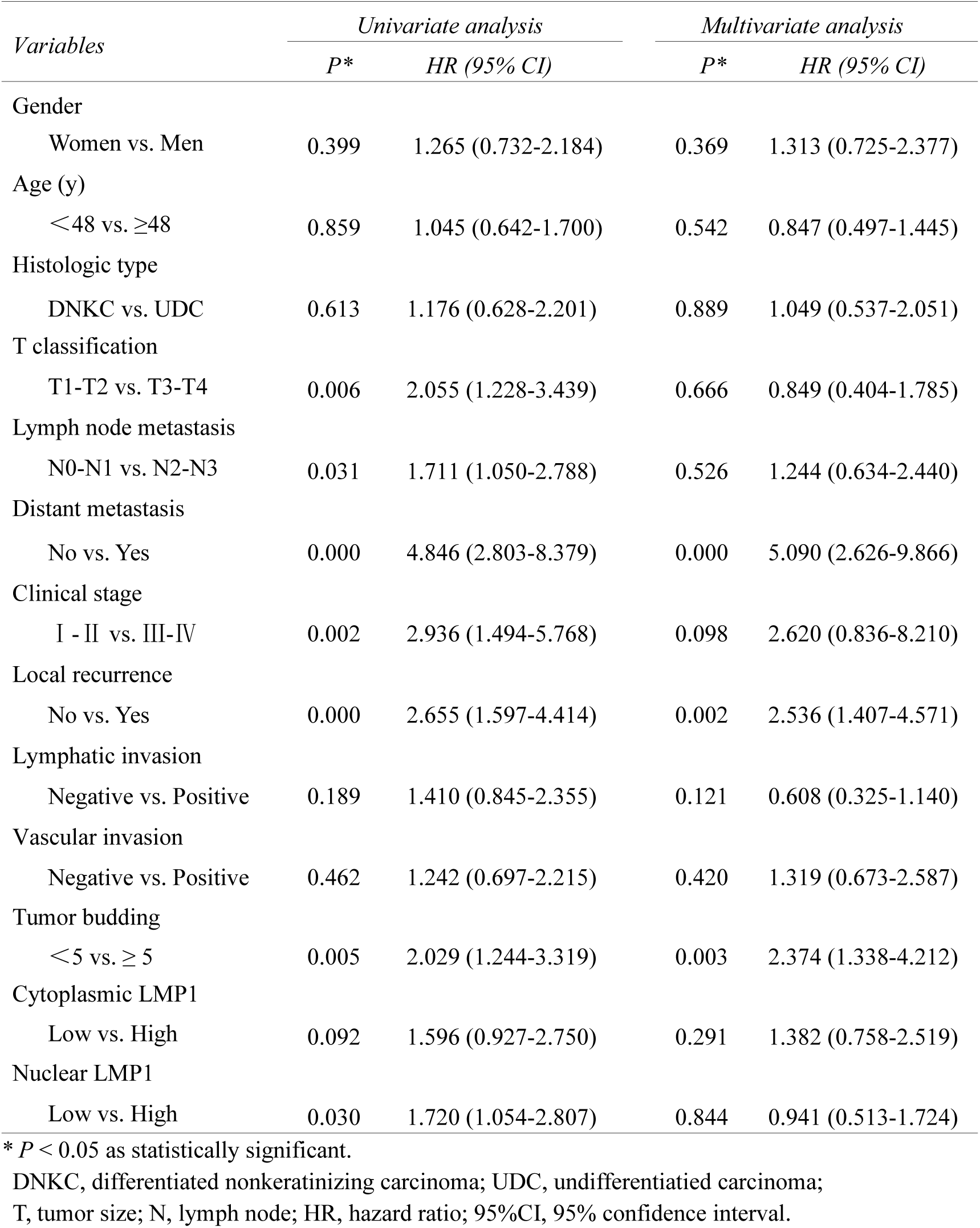
Cox proportional hazard models on overall survival of patients with NPC

## Discussion

With regard to the prognostic significance LMP1 in NPC, the issue is still controversial. For example, Hariwiyanto B et al. found that there was a significant association between LMP1 expression and overall survival of NPC patients (Hariwiyanto *et al*. 2010). In contrast, Chen CL et al. failed to find this link (Chen *et al*. 2006). In addition, some research groups described that it was not a valuable indicator of patients’ outcome, but linked significantly with clinicopathologic features including lymph node metastasis (Sarac *et al*. 2001; Ozyar *et al*. 2004). The discrepancy might be most likely due to tumor heterogeneity (including the different numbers and regions of samples) and protocol methods.

Firstly, we found that LMP1 protein was significantly elevated in 136 tumors compared with 45 non-neoplastic nasopharyngeal tissues, which is consistent with other previous observations (Hariwiyanto *et al*. 2010; Sarac *et al*. 2001; Taheri-Kadkhoda *et al*. 2009), indicating that EBV-relate proteins may play a critical role in the pathogenesis of NPC. Numerous studies have showed that LMP1 protein was detectable in the cytoplasm (or membrane) of cancer cells. In the present study, we observed that LMP1 was localized in both the cytoplasm and nucleus of tumor cells in NPC. Furthermore, a significant relationship was observed between cytoplasmic LMP1 and lymphatic invasion and vascular invasionin of NPC. On the other hand, high expression of nuclear LMP1 also linked tightly with aggressive aspects of tumors, including lymph node metastasis, local recurrence, lymphatic invasion. For example, 60.6% of patients with local recurrence displayed high levels of nuclear LMP1 in comparison with 30.1% of those without local recurrence, suggesting nuclear LMP1 overexpression might enhance the risk of tumor relapse, and might be a potent predictor of patient survival. As we know, local recurrence is the major cause of treatment failure and a worse prognosis in patients (Lee *et al*. 2012). Based on these findings, we suggest that aberrant expression of LMP1 might contribute to tumor aggressiveness and poor outcome of NPC. As anticipated, high nuclear LMP1 expression was significantly associated with shorter overall survival. However, similar to other research groups, no significant difference was found between increased expression of cytoplasmic LMP1 and patients’ survival (Li *et al*. 2009; Chen *et al*. 2006). To our knowledge, it is the first time to divide aberrant localization of LMP1 expression into individual variables of patient outcome.

Epithelial-mesenchymal transition (EMT) is responsible for tumor immunosurveillance and invasion, and contributes to poor outcome of various cancer types (Zidar *et al*. 2018; Horikawa *et al*. 2007). Recently, we have also reported that EMT might occur in the progression of NPC, and EMT-related factors E-cadherin and Vimentin were unfavorable prognostic factors in patients outcome (Luo *et al*. 2012). LMP1, which is the major EBV oncoprotein, has been proved to be sufficient to induce EMT in NPC cell lines. For example, Horikawa T et al demonstrated that forced expression of LMP1 could enhance cell migration and invasiveness, and activate the EMT program *in vitro* (Horikawa *et al*. 2011). Similarly, Shair KH et al. described that up-regulation of LMP1 could greatly induce a cadherin switch from E-to N-cadherin in NPC C666-1 cell lines (Shair *et al*. 2009). In 1998, we have shown that transfecting LMP1 promoted the invasion and metastasis of NPC by inhibiting E-cadherin expression (Ou *et al*. 2008). As to the relationship between LMP1 and EMT in NPC tissues, Horikawa T also found that LMP1 correlated significantly with E-cadherin in NPC samples. In this study, we found that both cytoplasmic and nuclear of LMP1 correlated inversely with expression of E-cadherin, whereas linked positively with expression of Vimentin, N-cadherin and Snail. Based on these observations, we conclude that EBV-encoded protein LMP1 might be crucial for the acquisition of aggressive and invasive properties in NPC.

The tumor invasive front was first discovered in colorectal cancer in the early 1980s, which is a prognostic area determining the aggressive behaviors and biological processes of cancers. Accumulating evidence has demonstrated that EMT is frequently present at the invasive front of tumors (Christofori. 2006; Liang. 2011). Of note, we have recently reported that aberrant expression of EMT-related molecules E-cadherin, Vimentin and Snail was predominantly observed at the invasive front of NPC (Luo *et al*. 2012b; Luo *et al*. 2012c). In this study, a preferential increase of nuclear LMP1 was also found in the tumor invasive front. We postulate that these tumor cells with high expression of nuclear LMP1 might be endowed with more invasive and aggressive potentials. As expected, we found that nuclear LMP1 overexpression in the invasive front correlated strongly with various aggressive features including T classification, lymph node metastasis, lymphatic invasion and local recurrence in NPC.

On the other hand, during the EMT process, single cells or small clusters of cancer cells along the invasive front of tumors, called “tumor budding”, are fragile to escape from the primary tumor. Tumor budding is a histological feature and has been observed in various human cancers including colorectal cancer and pancreatic cancer (Betge *et al*. 2012; Karamitopoulou *et al*. 2013). More importantly, it is recognized as a valuable prognostic predictor by UICC. In NPC specimens, we have also described the presence of tumor budding and its associations with aggressive behaviors and poor survival of patients (Luo *et al*. 2012d). Here, we show a distribution of nuclear LMP1 in tumor budding cells, and the protein patterns linked significantly with the degree of budding cells. Moreover, high expression of nuclear LMP1 was more frequently occurred in tumor cells invading into the surroundings, and these cells always exhibited a spindle-shaped morphology. Of note, our previous report had already found that malignant spindle cells correlated closely with EMT, and should be the more aggressive subtype in NPC (Luo *et al*. 2012a). Interesting, Ding RB et al has recently also showed that these sarcomatoid cells exhibited enriched EMT and invasion promoting genes (Ding *et al*. 2021). Taken together, our results imply that nuclear LMP1 may promote greatly the dynamic progression of NPC.

In conclusion, our work demonstrates the existence of nuclear LMP1 in NPC tissues. However, the functions of nuclear LMP1 in relation to NPC initiation and progression remain unknown. In addition, the molecular mechanisms regulating nuclear translocation of LMP1 also need to be clarified. LMP1 has been proved to play an important role in NPC pathogenesis, and we believe that elucidating these above issues might definitely provide novel therapeutic strategies of patients.

## Supporting information

Clinicopathologic features of 136 NPC tissues and EMT-related proteins expression

## Data Availability

All data produced in the present study are available upon reasonable request to the authors

## Abbreviations

LMP1: Latent membrane protein-1
NPC: nasopharyngeal carcinoma
EMT: epithelial-mesenchymal transition
TMA: tissue microarray
IHC: immunohistochemistry
FFPE: immunofluorescence labelling of formalin-fixed
DNKC: differentiated nonkeratinizing carcinoma
UDC: undifferentiatied carcinoma
T: tumor size
HR: hazard ratio
CI: confidence interval

## Acknowledgments

This work was supported by National Natural Science Foundation of China (Grant No. 81202125, 81872202); Natural Science Foundation of Guangdong Province of China (Grant No. 2018A030313778), Natural Science Foundation of Shenzhen (Grant No. JCYJ20190809154603583, JCYJ20210324131210030, JCYJ20200109115420720).

## ICMJE DISCLOSURE FORM

**Date:** <u>2/10/2022</u>

**Your Name:** <u>Weiren Luo</u>

**Manuscript Title:** <u>Nuclear and Cytoplasmic Expression of LMP1 Correlates with Epithelial-mesenchymal Transition in Nasopharyngeal Carcinoma</u>

**Manuscript Number (if known):** <u>Click or tap here to enter text.</u>

In the interest of transparency, we ask you to disclose all relationships/activities/interests listed below that are related to the content of your manuscript. “Related” means any relation with for-profit or not-for-profit third parties whose interests may be affected by the content of the manuscript. Disclosure represents a commitment to transparency and does not necessarily indicate a bias. If you are in doubt about whether to list a relationship/activity/interest, it is preferable that you do so.

The author’s relationships/activities/interests should be defined broadly. For example, if your manuscript pertains to the epidemiology of hypertension, you should declare all relationships with manufacturers of antihypertensive medication, even if that medication is not mentioned in the manuscript.

In item #1 below, report all support for the work reported in this manuscript without time limit. For all other items, the time frame for disclosure is the past 36 months.

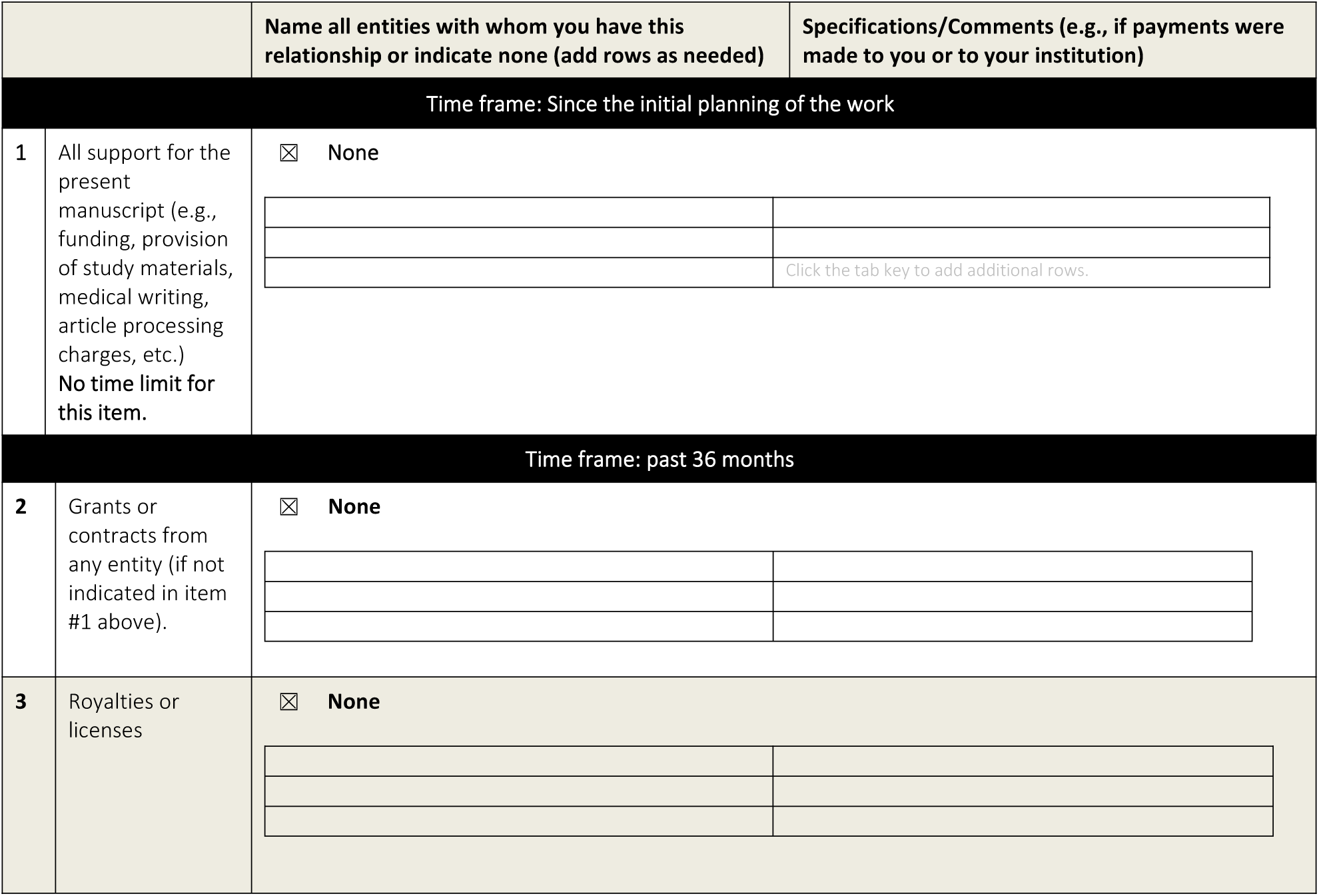
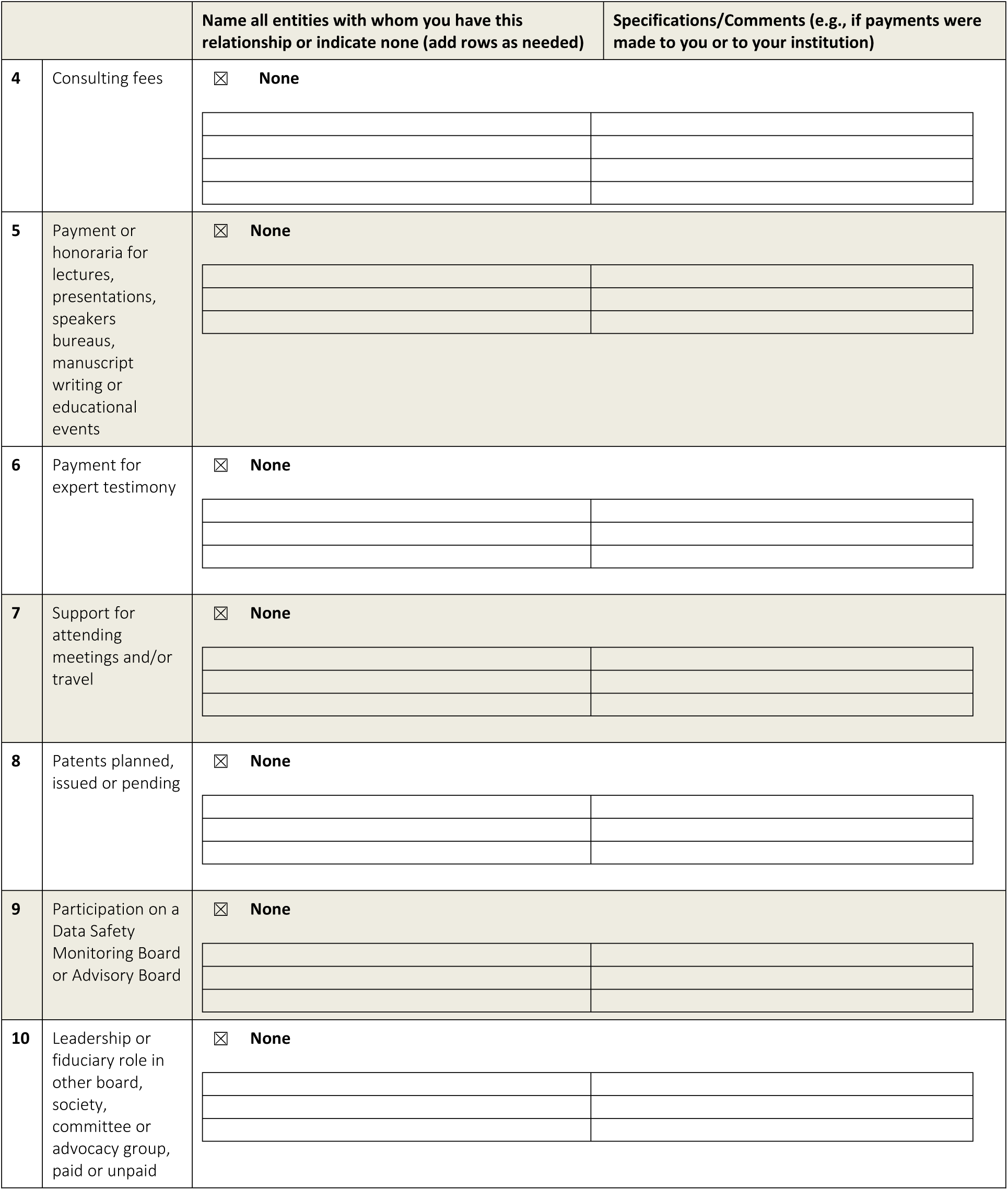
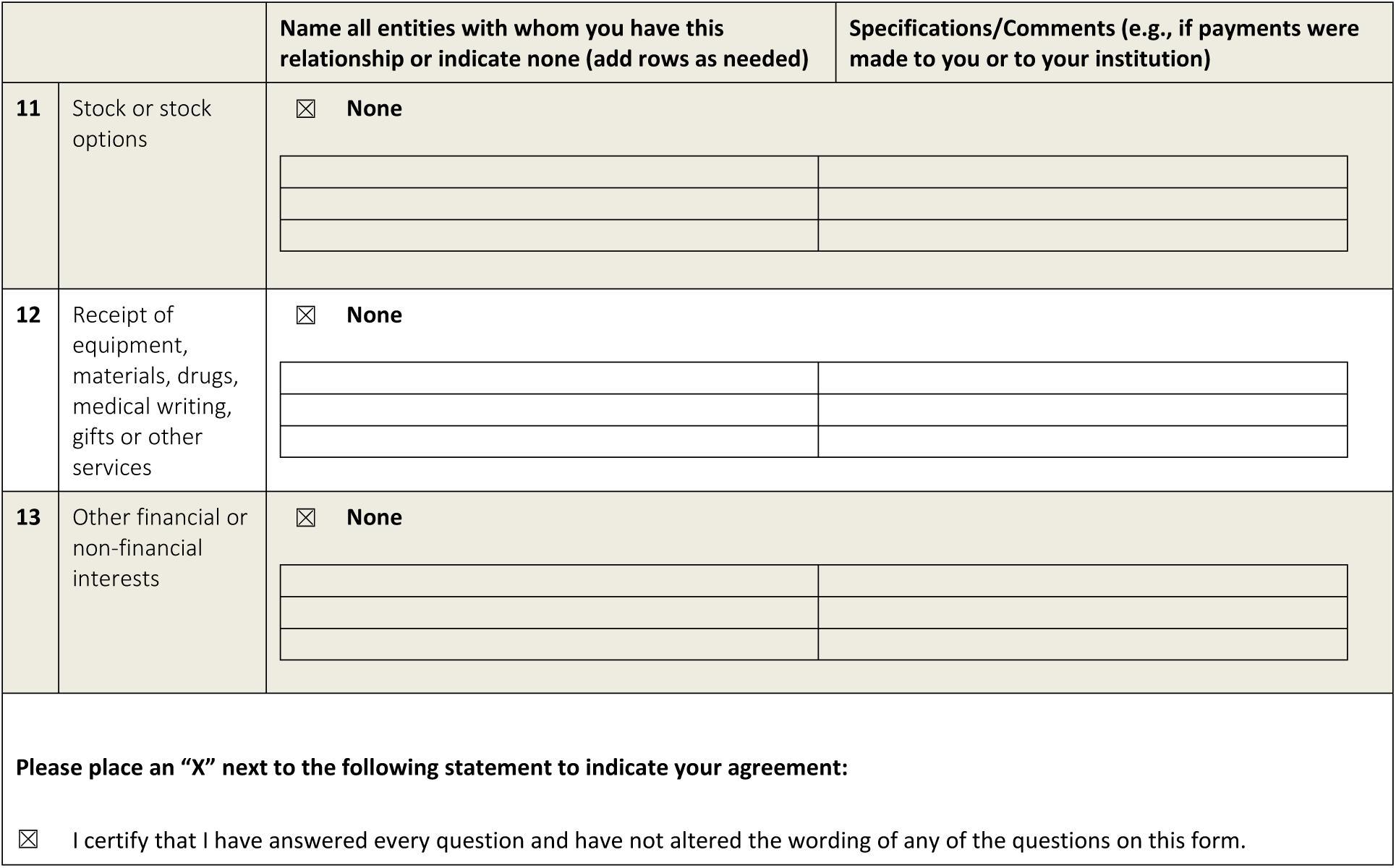

