## Supplementary material for "Cytoplasmic Expression of LMP1 and its Nuclear Translocation in Nasopharyngeal Carcinoma Correlates with Epithelial-mesenchymal Transition": Clinicopathologic features of 136 NPC tissues and EMT-related proteins expression

**Supplemental Table 1.** Clinicopathologic features of 136 NPC tissues and EMT-related proteins expression

| *Factors* | *n (%)* |
|---|---|
| Gender | |
| Women | 33 (24.3) |
| Men | 103 (75.7) |
| Age (y) | |
| ＜48 | 68 (50.0) |
| ≥48 | 68 (50.0) |
| Histologic type | |
| DNKC | 26 (19.1) |
| UDC | 110 (80.9) |
| T classification | |
| T1-T2 | 62 (45.6) |
| T3-T4 | 74 (54.4) |
| Lymph node metastasis | |
| N0-N1 | 80 (58.8) |
| N2-N3 | 56 (41.2) |
| Distant metastasis | |
| No | 115 (84.6) |
| Yes | 21 (15.4) |
| Clinical stage | |
| Ⅰ-Ⅱ | 38 (27.9) |
| Ⅲ-Ⅳ | 98 (72.1) |
| Local recurrence | |
| No | 103 (75.7) |
| Yes | 33 (24.3) |
| Lymphatic invasion | |

| | |
|---|---|
| Negative | 97 (71.3) |
| Positive | 39 (28.7) |
| Vascular invasion | |
| Negative | 108 (79.4) |
| Positive | 28 (20.6) |
| Tumor budding | |
| ＜5 | 80 (58.8) |
| ≥ 5 | 56 (41.2) |
| A total of LMP1 | |
| Low expression | 68 (50.0) |
| High expression | 68 (50.0) |
| Cytoplasmic LMP1 | |
| Low expression | 106 (77.9) |
| High expression | 30 (22.1) |
| Nuclear LMP1 | |
| Low expression | 85 (62.5) |
| High expression | 51 (37.5) |
| E-cadherin | |
| Low expression | 104 (76.5) |
| High expression | 32 (23.5) |
| Vimentin | |
| Low expression | 71 (52.2) |
| High expression | 65 (47.8) |
| N-cadherin | |
| Low expression | 73 (53.7) |
| High expression | 63 (46.3) |
| Fibronectin | |
| Low expression | 94 (69.1) |

| | |
|---|---|
| High expression | 42 (30.9) |
| Snail | |
| Low expression | 70 (51.5) |
| High expression | 66 (48.5) |
| Slug | |
| Low expression | 77 (56.6) |
| High expression | 59 (43.4) |
| Twist | |
| Low expression | 63 (46.3) |
| High expression | 73 (53.7) |

EMT, Epithelial-mesenchymal transition; DNKC, differentiated nonkeratinizing carcinoma; UDC, undifferentiatied carcinoma; T, tumor size
